# Automatic Identification of Tinnitus Malingering Based on Overt and Covert Behavioral Responses During Psychoacoustic Testing

**DOI:** 10.1101/2022.01.25.22269837

**Authors:** Christopher J. Smalt, Jenna A. Sugai, Elouise Koops, Kelly N. Jahn, Kenneth E. Hancock, Daniel B. Polley

## Abstract

Tinnitus, or ringing in the ears, is a prevalent condition that imposes a substantial health and financial burden on the patient and to society. The diagnosis of tinnitus, like pain, relies on patient self-report. Subjective self-report measures can complicate the distinction between actual and fraudulent claims and obscure accurate severity assessments. In this study, we combined tablet-based self-directed hearing assessments with neural network classifiers to objectively determine tinnitus severity, and to differentiate participants with tinnitus (N=24) from a malingering cohort, who were instructed to feign an imagined tinnitus percept (N=28). We identified clear differences between the groups, both in their overt rating of tinnitus severity but also covert differences in their fingertip movement trajectories on the tablet surface as they performed the reporting assay. Using only 10 minutes of data, we achieved 81% accuracy classifying patients vs malingerers (ROC AUC=0.88) with leave-one-participant-out cross validation. Objective measurements of tinnitus will improve estimates of tinnitus prevalence and help to prioritize and direct funds for tinnitus compensation.

## 1 Introduction

Tinnitus is a prevalent auditory condition that imposes a substantial burden to the patient and to society (Maes et al., 2013; Bhatt et al., 2016). According to the US veteran benefits administration fiscal year 2020, tinnitus was the most prevalent service-connected disability among new compensation recipients. In fiscal year 2020, 10.2% of all new benefits recipients received compensation for tinnitus (149,368 claims). Tinnitus was also the most prevalent overall disability with a total of 2,327,387 claims of tinnitus that year, with hearing loss as the second most prevalent disability claimed, representing 1,343,013 claims. The economic and health impacts of tinnitus are alarming not only because of the high prevalence, but also because tinnitus claims have been consistently rising at an average annual rate of 12% since 2011. It is noteworthy that since that time, claims have been growing at nearly twice the rate of hearing loss (7%), a related condition for which there is an established diagnostic measurement via the pure tone audiogram.

Currently, the diagnosis of tinnitus, like pain, relies on subjective and self-reported measures (Basile et al., 2013). As a result, it is challenging for patients to convey the characteristics and severity of their tinnitus percept to their caregivers. Although psychoacoustic measures such as tinnitus pitch and intensity matching are often obtained to define the auditory attributes of tinnitus, there is presently no established relationship between these attributes and the actual severity of the symptom(Henry, 2016; Byun et al., 2010). Both the quality and severity of tinnitus can vary over time for an individual (Henry & Meikle, 2000; Chen et al., 2020). Test-retest reliability across patients has been shown to be as high as 0.94 on a 52-item questionnaire (Hiller et al., 1994), but much poorer across longer time spans of months (Hoare et al., 2014) reflecting the dynamic nature of tinnitus.

Developing new, more objective, tinnitus diagnostics could prove useful for assessing tinnitus severity and for distinguishing been legitimate and fraudulent cases among the large - and ever growing - tinnitus disability claims. In this context, malingering is the feigning of a medical condition for gain. Possible incentives for doing so include obtaining economic compensation or to be removed from difficult circumstances, such as military service (Jerger et al., 1981). Previous work has shown mixed results in distinguishing malingerers from tinnitus patients using psychoacoustic measures (Henry, McMillan, et al., 2013; Steiger et al., 2013), partially due to the lack of reliability within a given patient with tinnitus (McMillan et al., 2014). In this study we develop a system that automatically classifies (i.e. “identifies”) tinnitus patients from malingerers through behavioral testing that establishes a relationship between the characterization of tinnitus loudness and acoustic properties and the presence of the disability.

## 2 Methods

### 2.1 Populations

A population of 52 participants were included for study. The tinnitus cohort (N=24) were recruited from Mass Eye and Ear, having reported tinnitus as their chief complaint, as reported in our previous publication (Chen et al., 2020). The malingering cohort consisted of 28 individuals who confirmed having no perception of tinnitus. This study was approved by the Massachusetts Eye and Ear Institutional Review Board and participants provided written informed consent to take part in the study. Participants completed several tablet-based tasks related to their hearing with calibrated headphones over a period of days to behaviorally characterize their tinnitus. Figure 1(A) schematizes the overview of the study design, where five sessions were performed over a 2 week period following a baseline clinical assessment and audiometry. After first confirming that they did not have chronic subjective tinnitus, participants in the malingering groups were instructed to perform all measures as if they heard a constant phantom sound. They were first trained on what tinnitus sounded like by reading text descriptions and listening to five sample audio files of tinnitus match sounds. Matched sounds were generated from data corresponding to actual patient matches of their tinnitus precepts (Chen et al., 2020). After confirming that they were confident in their ability to imagine a tinnitus sound, the malingering participants were instructed to complete the tests while imagining the presence of a constant tinnitus-like sound. Apart from these additional instructions and guidance for the malingering group, all procedures were matched between the two groups.

**Figure 1:**
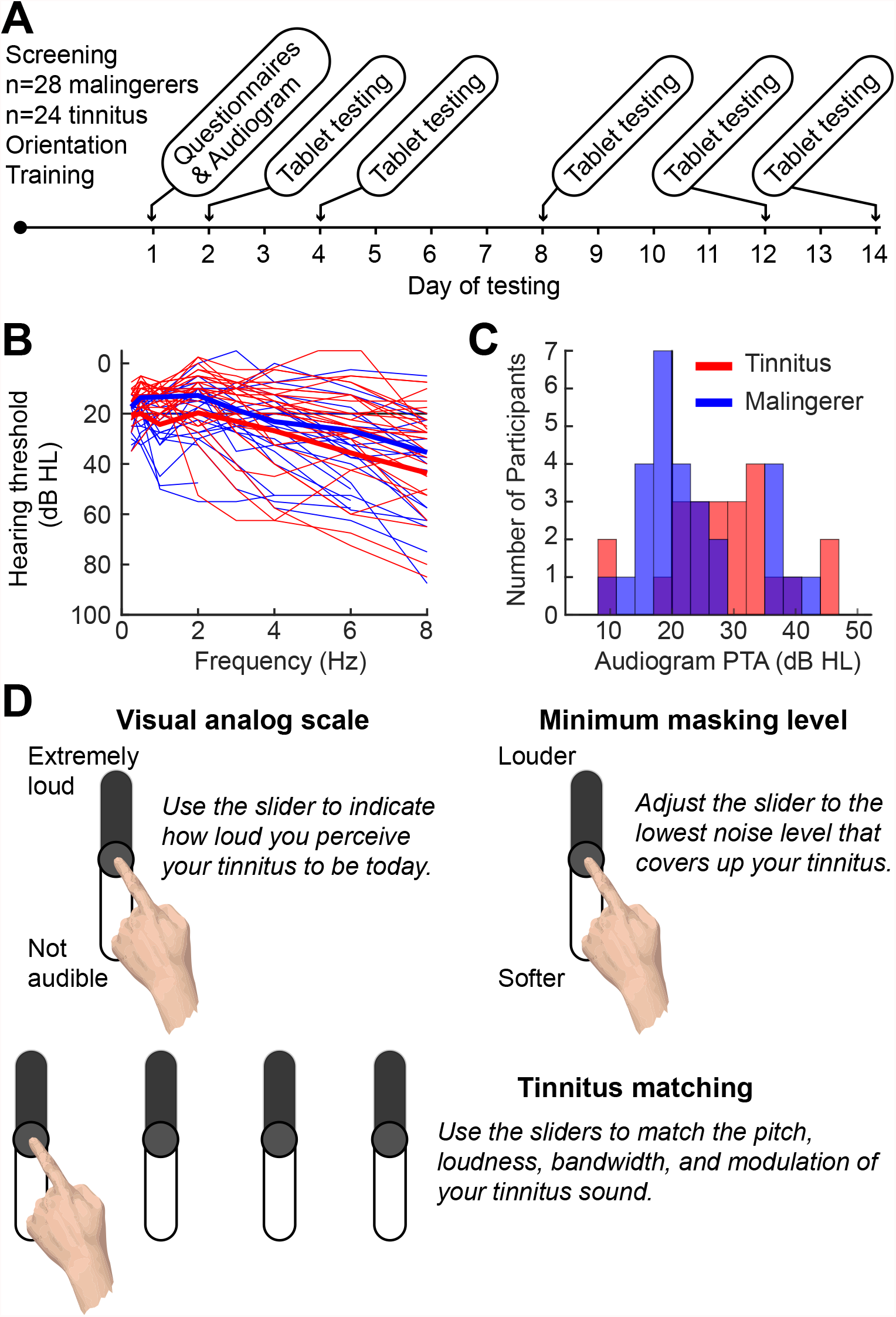
A-B: Audiograms for tinnitus patients (red) and malingerers (blue) who were instructed to pretend they had tinnitus during the remainder of the study. C-E: Graphical User Interface to characterize tinnitus loudness (D), minimum masking level (MML) (D) and to match tinnitus (E) with visual sliders. The overall order to the study is shown in F, where tinnitus matching was done in five separate sessions across a two week period using a tablet computer.

The average ages of the patient and malingerer groups were 52.0 and 53.7, respectively, with standard deviations of 12.3 and 11.4 respectively. Hearing (i.e audiometric) thresholds were measured for both ears between 0.25 and 16 kHz. Figure 1 panels B-C shows the individual and average audiograms for the two groups, respectively. This study was approved by the Massachusetts Eye and Ear and Mass General Brigham Institutional Review Boards.

### 2.2 Tinnitus characterization

All participants performed three psychoacoustic tasks: a tinnitus visual analog scale (VAS) rating, the minimum masking level (MML), and the tinnitus acoustic matching, all of which used a slider-bar and touch screen to collect the participant’s response. For each of the five experimental sessions, multiple runs (i.e. repetitions) were performed for each measurement type. Each of these repetitions within a session are referred to as trials. Subjects performed all testing from home using calibrated circumaural headphones (Bose) and custom software applications developed as a Windows Store App using the Unity game engine and side-loaded onto the tablets (Microsoft Surface Pro 2).

Figure 1D depicts the participant view of the three slider tasks and their instructions. The VAS slider ranged from ‘not audible’ to ‘extremely loud’ (0-100). In the MML task, participants adjusted a virtual slider to control the sound level of a noise band. Their task was to identify the minimum sound level at which they no longer perceived their (imagined) tinnitus over the masking noise. Finally, participants were then asked to adjust the acoustic properties of sound delivered to one ear so as to match the sound of their real or imagined tinnitus percept. The sound was presented monaurally, to the ear where participants reported having less tinnitus, thus allowing them to compare the sound generated by the tablet software to their perceived or imagined tinnitus sound. Subjects adjusted four auditory characteristics: level (dB SL), center frequency (Hz), bandwidth (octaves), and amplitude modulation (Hz). They could adapt these characteristics with sliders on the tablet via interaction with the touchscreen, where the sound could be changed procedurally as they adjusted the sliders. The parameters used to generate the audio were sampled and stored at approximately 5 times per second (5 Hz) from the slider values. The range of values the slider encoded became smaller over trials within a session. Thus, the same distance (i.e., the values covered, for instance, a change in level) a finger traveled on the slider in trial 1 does not necessarily correspond to the change in level in trial 10.

### 2.3 Feature Extraction

Two approaches were used to analyze the participant’s interaction with the graphical touch sliders, corresponding to *what* was measured and *how* it was answered. The first approach focused on the response outcomes, which correspond to standard pyschoacoustic measures of tinnitus including the masking level, VAS and tinnitus matching. These standard measures are derived from the final value of the graphical slider on each trial of the experiment. A repeated measures ANOVA with the factors session (1-5) and group (patient, malingerer) was then used to determine whether there were any differences between the reported precept of the two populations, as well as changes in test-retest reliability. Participants were modeled as a random effect. The second method focused on how participants interacted with the graphical user interface and sliders over time, not just the final value of the slider. Figure 2A-B show how the change in slider value over time can be recorded and stored as a time series.

**Figure 2:**
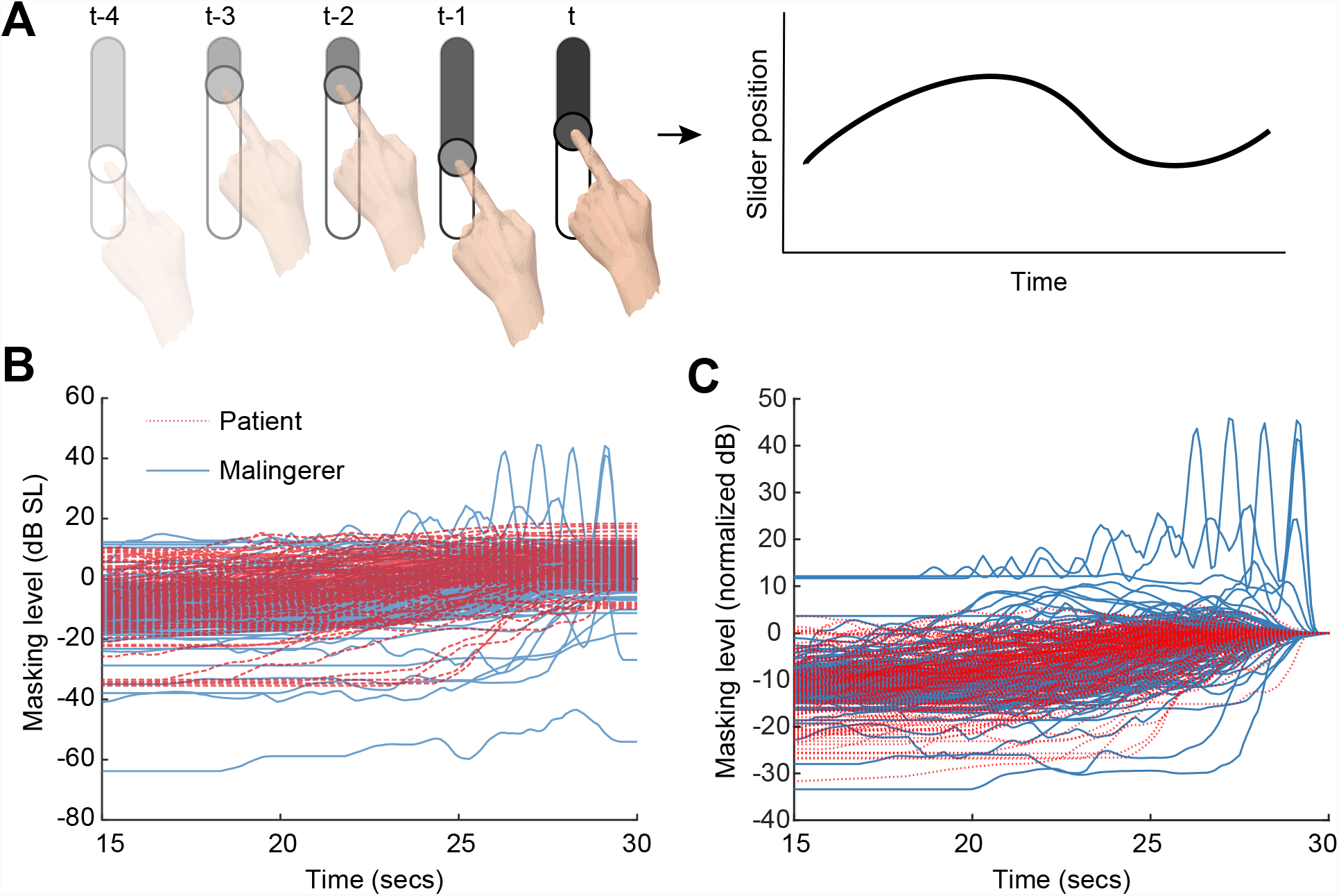
Minimum Masking Level (MML) slider position as a function of time for tinnitus patients and malingerers. Only the last 15 seconds of the track were extracted for further analysis.

### 2.4 Machine learning

Classifier algorithms were constructed to distinguish between tinnitus and malingerer groups. Leave-one-subject-out (LOSO) cross-validation was used to train and evaluate the model, combining runs of slider adjustment across the various tinnitus-characterization tasks within an experimental session. Classifiers were evaluated in terms of classification accuracy (percent correct), and by the area under the curve (AUC) of the receiver operating characteristic (ROC) curve, which evaluates the diagnostic capability of a binary classifier (Fawcett, 2006).

Classification was performed with three different types of input data and corresponding classifiers. First the outcome measurements alone were used (i.e. the final slider values) as input using (multiple) logistic regression. Second, the MML slider time series was used as input to a convolutional deep neural network (DNN) to investigate latent information stored in the slider movement. Finally, summary features were derived and pooled from the time-series data of all tasks. Feature-based classifier implementation was done in Python using scikit-learn’s `LogisticRegression` and `RandomForestClassifier` (Pedregosa et al., 2011) in a leave-one-participant out fashion. The random forest classifier was run with with 100 estimators and a maximum depth of 5 trees. Features derived included the slider final value, the mean and standard deviation of the slider time-series across runs as well as the maximum velocity, and cross-correlation of the time-series across runs.

Alternatively, a DNN was applied to the raw slider time series data derived from the tinnitus masking task that did not depend on manual feature selection. Because the time-series may be a different length for each participant and each trial, only the final 15 seconds of slider time-series for each participant was used (see Figure 2 C). This fixed duration segmentation also had the benefit of removing time spent on each trial as a latent feature (i.e. participant effort).

The DNN was implemented in pytorch, and used two cascaded 1D convolutional layers (18 and 9 channels respectively), followed by maxpool, batchnorm, and a fully connected layer. Eighteen input channels corresponded to the 18 trials of the MML task performed in a single session. This means that there were a total of five sessions evaluated by the DNN for each participant. Dropout was used during training with a probability of 10%.

To separate *how* the participant interacted with the slider from *what* the final result of the slider revealed (i.e. the MML) we subtracted the mean slider final value from the time series for each participant. This process normalizes the actual selected behavioral value so that the final value is 0 dB SL on average (Figure 2).

## 3 Results

### 3.1 Tinnitus Characterization

Patients and malingerers quantified their perceived severity and psychoacoustic qualities of their (imagined) tinnitus in several ways, including visual analog scale rating, minimum masking level (MML), and sound matching. Figure 3 illustrates the MML (A), the visual analog scale (VAS) tinnitus rating (B), as well as the sound level (C) and spectral bandwidth (D) that matches the (imagined) tinnitus sound. When compared to patients with tinnitus, malingerers reported that their tinnitus was masked at lower sound levels, was less bothersome, and had broader spectral bandwidth (i.e., less like a pure tone) (Repeated-measures ANOVA main effect for group on MML (*F* = 9.28, *p* = 0.004), VAS rating (*F* = 31.7, *p <* 0.0001), matching bandwidth (*F* = 7.31, *p* = 0.009). No differences were noted in the sound level used to match the tinnitus loudness between groups (*F* = 0.005, *p* = 0.94). No significant effect of experimental session or interaction effect was observed.

**Figure 3:**
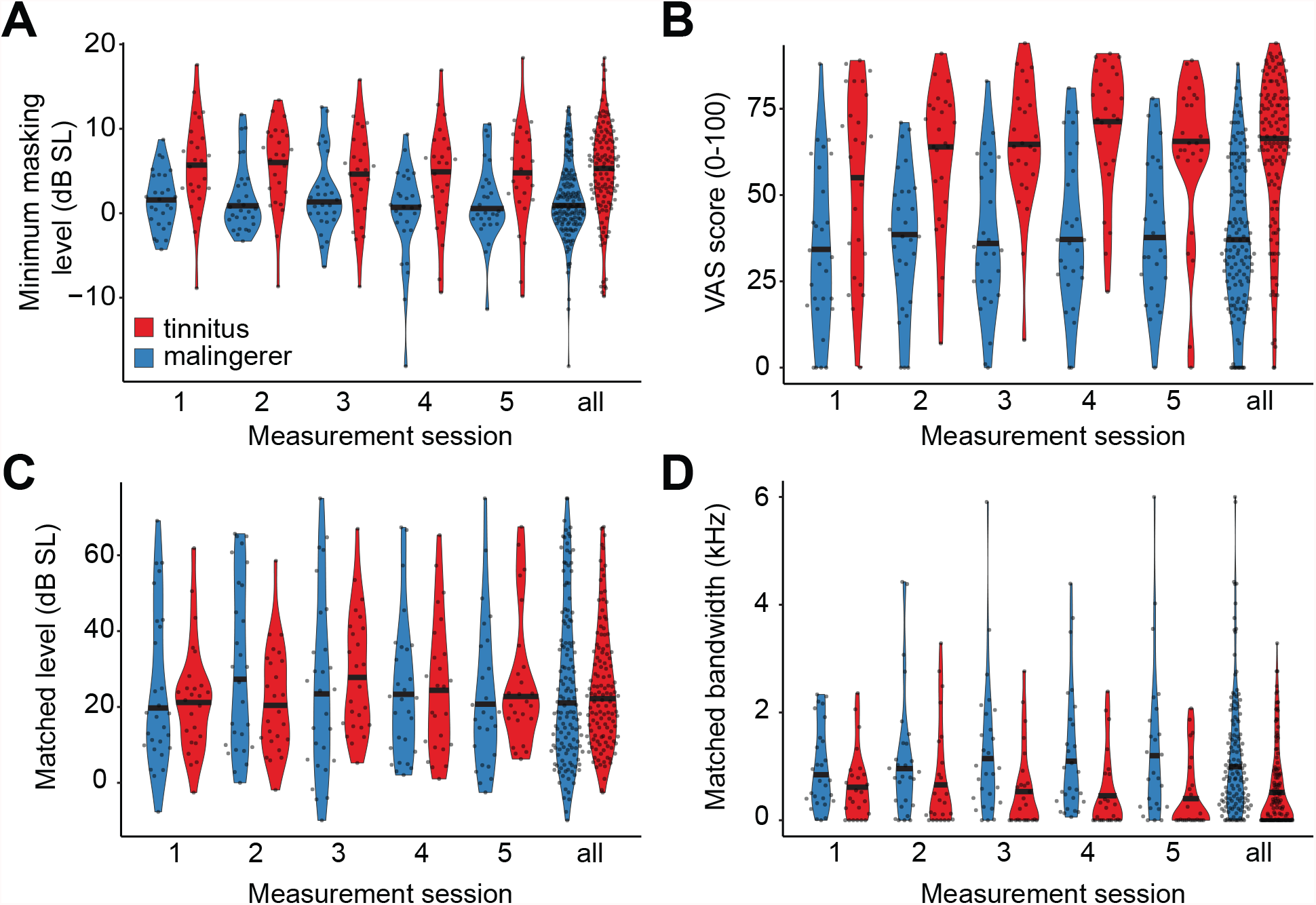
In our sample population, malingering participants report that their imagined tinnitus is less tonal, more easily masked, and less bothersome than patients with tinnitus. Normalized density distributions along with single participant data (filled circles) are provided for MML (A), VAS (B), loudness matching (C) and bandwidth matching (D). All estimates were repeated five times, shown individually and collapsed over all sessions.

### 3.2 Classification based on outcome measures

While pyschoacoustic and self-reported tinnitus measures indicate statistical differences between the tinnitus patient and malinger groups, our motivation for the study was to determine if tinnitus status could be determined objectively on the individual patient level. Figure 4 shows how accurately logistic regression models were able to classify patients from malingerers considering only the final value of each slider (the outcome variable(s)) for each task as the predictor. The MML and VAS classifiers were trained using a single outcome variable corresponding to individual sliders, while the classifier for the matching task had four corresponding to the four sliders used. Finally, the performance combining all slider outcomes is shown in red, achieving 68% Accuracy and an AUC=0.75.

**Figure 4:**
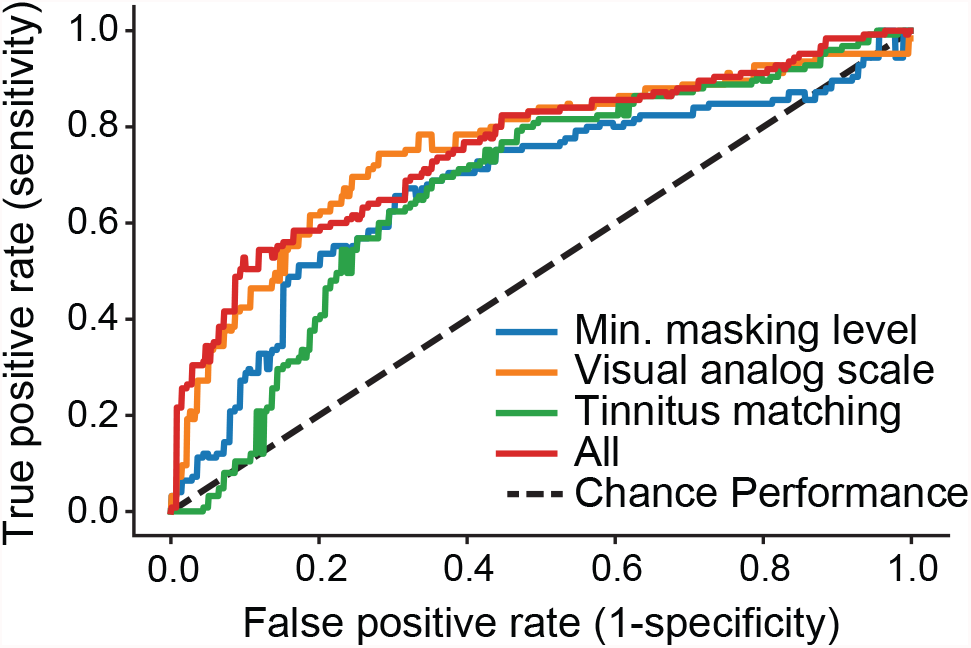
Receiver operator characteristic linear regression on tinnitus psychoacoustic task outcomes only. Combining the outcomes across MML, VAS, and Matching tasks achieved a 66% Accuracy and an AUC=0.75

### 3.3 Slider time-series

In addition to classifying patients from malingers based on the final outcome variables (the final placement of each individual slider), we also created a model to classify the two groups based on engagement of the participant with the sliders during the entirety of the MML task. Figure 5A compares the approach using only the pyschoacoustic outcome variable (logistic regression, green) with two alternative classification approaches using the additional features derived from the slider time-series. The feature-based random forest (orange) and DNN (blue) utilize the raw slider time series shown in Figure 2C. The respective performances were AUC=0.67, 0.77, and 0.84 with corresponding accuracies of 49%, 65%, and 77%. The Random Forest, which incorporates features such as participant variability within a session, outperforms the classifier relying on the outcome variable alone (logistic regression). The DNN outperforms both with the highest performance, likely due to the access to raw data and potential latent features in the data.

**Figure 5:**
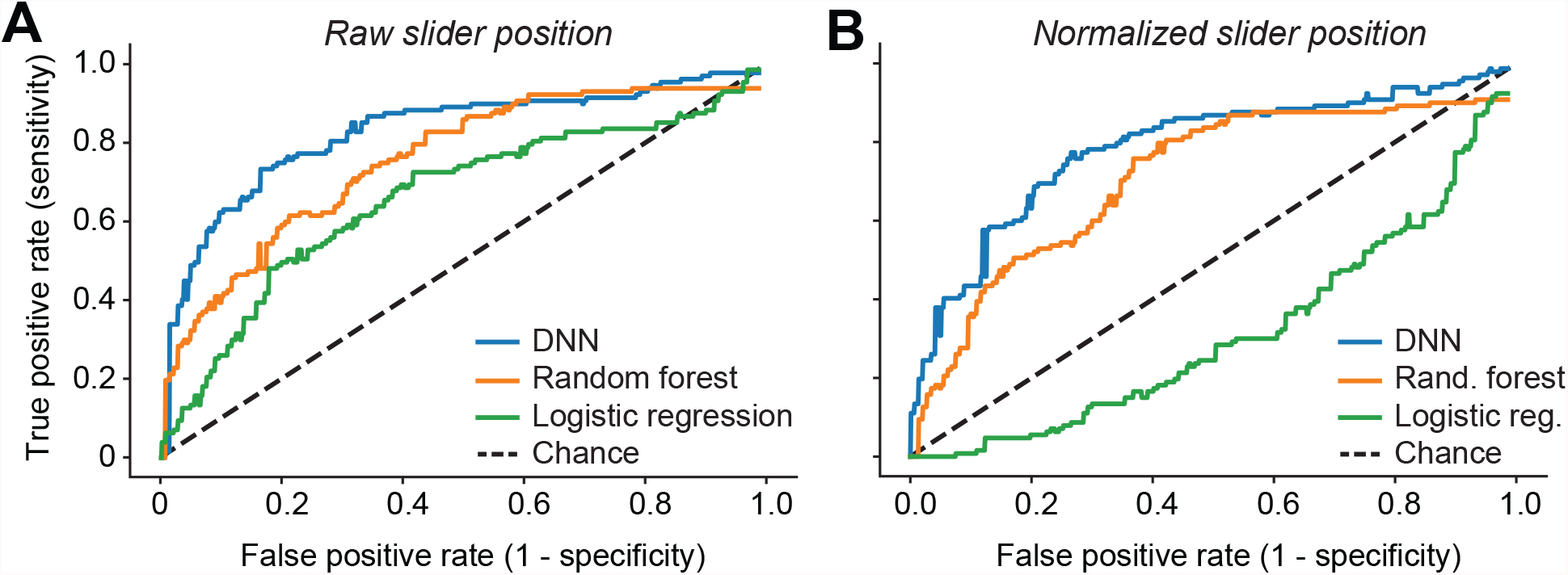
Patient vs malingerer classifier performance using minimum masking level (MML) slider time-series data. Panel A uses the original time-series data with ROC AUC=0.84, 0.77, and 0.67 and accuracies of 77%, 65%, 49% accuracy respectively for a DNN, Random Forest, and Logistic Regression. Panel B ran the same classifiers on time-series data where the MML value is subtracted out for each participant with an ROC AUC=0.79, 0.72, and 0.31 and 77.4%, 65%, 49% accuracy respectively.

A close inspection of the finger path trajectories on the MML task revealed that tinnitus participants interfaced with the virtual slider differently than malingering participants. As illustrated in (Fig. 2), tinnitus participants gradually increased the making level to converge on their MML, whereas malingering participants made larger, more erratic adjustments before abruptly stopping on their MML. To analyze the degree that the DNN relied on the slider time-series versus the outcome variable (MML), we subtracted the MML for each subject from each time-series (Figure 2D). This normalization resulted in the respective ROC: AUC=0.79, 0.72, and 0.31, and accuracies of 77.4%, 65%, 49% for the DNN, random forest, and logistic regression, respectively (Figure 5B). As expected, the logistic regression performed at chance, since it only used the outcome variable that had been normalized away on average. However, the other two methods show only a slight dip in performance, indicating that the manner in which the slider was adjusted, i.e., how malingering subjects adjust the slider over time rather than the final psychoacoustic measurement value, can be used to discriminate the two populations.

### 3.4 Feature-based classification

Finally, we considered a feature-based approach that captured both the outcome variable and slider interaction across all three tasks (VAS, MML, Matching) while maintaining interpretability (as opposed to a Neural Network). ROC curves are shown on each task separately and combined in a single system (Figure 6A). The MML, VAS, and Matching tasks separately achieved AUCs of 0.68, 0.72, and 0.70 and accuracies of 65%, 70%, 66%, respectively. The combined system performance additionally included other demographic information, such as age and sex, as well as the subjective tinnitus handicap inventory (THI) score. The combined system performance achieved 81% accuracy with an ROC of 0.88, outperforming any individual task.

**Figure 6:**
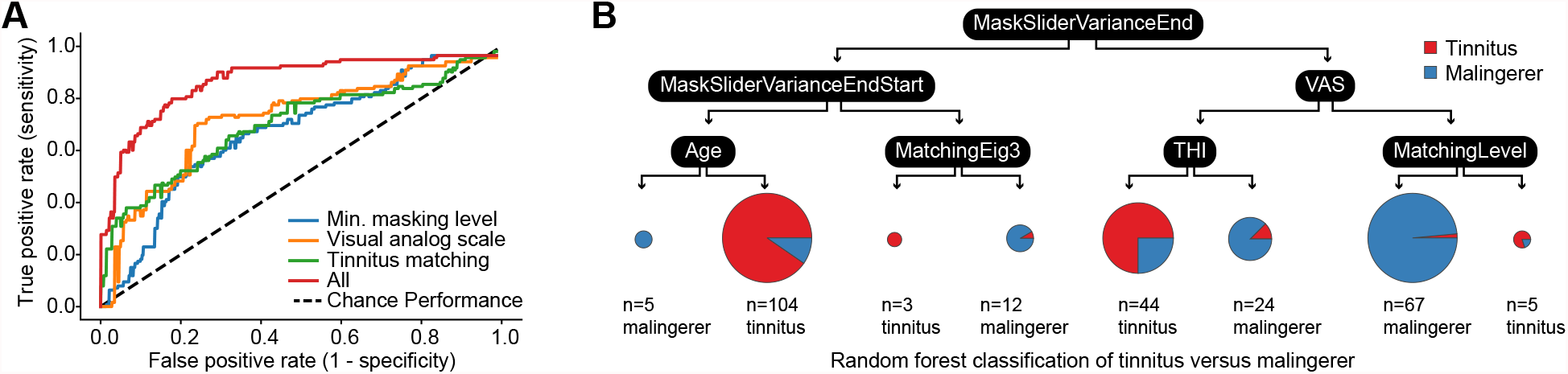
ROC for tinnitus versus malingerer classification on three psychoacoustic tasks (blue, green, yellow), and combined for all tasks (red) using a random forest. Combined performance accuracy was 81% (AUC = 0.88).

Figure 6B illustrates a single decision tree from the random forest derived from features across all three tasks. In this example tree, the root node feature is the standard deviation across trials of the MML slider end point (MaskingSliderStdTrackEnd). Typically, the root node feature is best at separating the two populations, while other features further down in the tree are important but of decreasing use in the separation; in this example, these features include the VAS slider value, the subject age and THI. An advantage to this type of classifier is in its interpretability for medical diagnoses or financial decisions.

## 4 Discussion

This study aimed to distinguish participants with tinnitus from participants feigning tinnitus using their responses on several standard tinnitus characterization tests. Our results demonstrate that it is possible to discriminate individuals with tinnitus from malingerers with an accuracy of 81% using approximately 10 minutes worth of reporting data. On average, tinnitus patients reported a higher MML relative to their audiometric threshold, and higher loudness on a visual analog scale and through tinnitus matching. By contrast to prior reports, we did not observe that malingering participants matched their imagined tinnitus to higher physical sound levels than participants with tinnitus (Byun et al., 2010).

However, in this study we went beyond characterizing the group differences and extracted information about how the participant interacts with their tablet as opposed to only utilizing their psychoacoustic test results. Our analysis revealed that information about the populations is contained in both how they interacted with the slider as well as in the final slider value. This result has implications for many other fields of subjective or neuropsychiatric research (e.g. pain), where consumer-grade electronics are increasingly being used (Yang et al., 2019; Chen et al., 2020; Henry, Roberts, et al., 2013).

Variation across trials within a session proved useful for classifying participants as patients or malingers (see Figure 6B). With chronic subjective tinnitus, the phantom sound percept is continuous but the perceptual qualities (the loudness, pitch, etc.) can fluctuate over time (Chen et al., 2020). For this reason, we included the natural heterogeneity within a participant by compiling measurements performed in five separate test sessions across several weeks. Condensing the measurements into a single session would reduce the variability within participants and presumably lead to even more accurate classification accuracy. This prediction could be tested in future studies but, for the purposes of this study, our classification accuracy and measurement time represents a conservative estimate.

On the individual trial level, patients feigning tinnitus took less time to mask their tinnitus and had fewer reversals while matching their tinnitus. This result is in line with the expectation that it takes more time to achieve a precise match or masking of actual tinnitus than of feigned tinnitus. However, we opted to not include this as a feature in the classification and relied on a subset of the raw time-series data. The primary reason for this is that the duration could be easily adapted and would not necessarily be a robust, repeatable measure and more likely reflects participant engagement.

The study is limited in several ways. While the samples were matched in terms of age, they were relatively small, and not controlled based on familiarity with using the tablet, the time of day, or audiometry. Also, the malingering participants in the current study were not financially motivated to feign tinnitus to the same extent as actual malingers, whose financial renumeration could be larger but also condition upon making a credible claim. Because of the simplicity of the tests reported and the minimal amount of patient time required to conduct them, more robust classifiers could be developed on larger datasets through online testing. A larger study could also be validated against a true held-out dataset rather than rely on cross-validation.

## 5 Conclusion

Conventional subjective tinnitus assessments based on questionnaires cannot easily detect malingerers. In the broader context of tinnitus clinical research, questionnaire assessments tend to inflate placebo effects, underestimate treatment effects, and generally obscure the true prevalence and severity of tinnitus in the population. While objective biomarker measurements are the gold standard and the ultimate objective for tinnitus diagnostics, automatic and quantitative assessments of biobehavioral data represent an important intermediate step. Further to this point, as studies are progressively moving to online formats, these data can be scaled up to larger cohorts and subjected to similar forms of classification analyses described here. It seems likely that automated analyses of standardized rapid tinnitus tests would inform the decision as to whether to accept or deny claims of tinnitus disability. In this way, accurate identification of participants with tinnitus and differentiation from malingerers will also help funds be distributed where they are needed most and enable research on tinnitus severity.

## Data Availability

All data are available from the corresponding author on reasonable request

## 6 Data Availability

The data are available from the corresponding author on reasonable request.

## 7 Competing Interests

The authors declare that there are no competing interests.

## 8 Author Contributions

C.S performed the data analysis, machine learning, and drafted the manuscript. E.H also contributed to the data analysis and writing of the manuscript draft. K.H wrote the real-time tablet software for data collection and storage. J.S. and K.J interacted with participants, and assisted with the study design. D.P. provided guidance in study design, analysis and writing. All authors reviewed the manuscript.

## 9 Acknowledgements

Address correspondence and reprint requests to Prof. Daniel Polley, 243 Charles Street, Boston, MA 02114; E-mail

The authors would like to thank Michael Brandstein for his review of the classifier design and implementation.

DISTRIBUTION STATEMENT A. Approved for public release. Distribution is unlimited.

This material is based upon work supported by the United States Air Force under Air Force Contract No. FA8702-15-D-0001. Any opinions, findings, conclusions or recommendations expressed in this material are those of the author(s) and do not necessarily reflect the views of the United States Air Force.

This work was supported by a research award from the Boston One Fund and NIDCD P50 DC015817.

## Notes

### Competing Interest Statement

The authors have declared no competing interest.

### Funding Statement

DISTRIBUTION STATEMENT A. Approved for public release. Distribution is unlimited.
\\ \\
\noindent This material is based upon work supported by the United States Air Force under Air Force Contract No. FA8702-15-D-0001. Any opinions, findings, conclusions or recommendations expressed in this material are those of the author(s) and do not necessarily reflect the views of the United States Air Force.
\\ \\
This work was supported by a research award from the Boston One Fund and NIDCD P50 DC015817.

### Author Declarations

IRB of Massachusetts Eye and Ear gave ethical approval for this work

